# Bayesian Logical Neural Networks for Human-Centered Applications in Medicine

**DOI:** 10.1101/2021.11.15.21266351

**Authors:** Juan G. Diaz O., Lukas Maier, Orsolya Csiszár

## Abstract

Medicine is characterized by its inherent ambiguity, i.e., the difficulty to identify and obtain exact outcomes from available data. Regarding this problem, electronic Health Records (EHRs) aim to avoid imprecisions in the data recording, for instance by its recording in an automatic way or by the integration of data that is both readable by humans and machines. However, the inherent biology and physiological processes introduce a constant epistemic uncertainty, which has a deep implication in the way the condition of the patients is estimated. For instance, for some patients, it is not possible to speak about an exact diagnosis, but about the “suspicion” of a disease, which reveals that the medical practice is often ambiguous. In this work, we report a novel modeling methodology combining explainable models, defined on Logic Neural Networks (LONNs), and Bayesian Networks (BN) that deliver ambiguous outcomes, for instance, medical procedures (Therapy Keys (TK)), depending on the uncertainty of observed data. If epistemic uncertainty is generated from the underlying physiology, the model delivers exact or ambiguous results depending on the individual parameters of each patient. Thus, our model does not aim to assist the customer by providing exact results but is a user-centered solution that informs the customer when a given recommendation, in this case, a therapy, is uncertain and must be carefully evaluated by the customer, implying that the final customer must be a professional who will not fully rely on automatic recommendations. This novel methodology has been tested on a database for patients with heart insufficiency.

## Introduction

In daily business, physicians are confronted with the constant integration and evaluation of different parameters to assess the patient’s condition and in this way, establish correct diagnoses as well as therapies, encoded as Therapy Keys (TKs). This assessment is particularly difficult for health professionals due to the constant work overload in health centers, as well as the cognitive impairment that represents selecting an item, like a potential therapy, from a portfolio with several options (Hall and Walton, 2004). To this end, support systems represent the best option to assist the daily work of health professionals; but these systems require mathematical models.

Traditionally such support systems have been defined as expert systems (Sandell and Bourne, 1985; Zhou and Sordo, 2021). Recently, with the development of efficient computing methods, the use of deep learning methods has found more acceptance, for instance for pattern recognition, extraction in EHRs and integration of the output to recommender systems (Rajkomar et al., 2018). However, the use of any model in medicine must fulfill the condition that this model should be explainable, i.e., the way how the results are obtained must be understandable by the customer.

Conventional methods from machine learning, like decision trees, are essentially explainable since the derivation of the final result can be tracked in the entire computation process. However, these methods are limited by their accuracy and its scalability, i.e., their ability to handle an ever-growing amount of information. To this end, deep learning methods are an attractive option over other modeling alternatives. Unfortunately, models based on this method are unexplainable, since the computation of the network’s weight’s obey internal coupled optimization processes that are difficult to be explained and presented to customers with a poor technical background in the field. To this end, great effort has been made in order to establish and standardize explainability in deep learning (Linardatos et al., 2020).

Alternatively, deep-learning models can be explainable with a change of the network’s structure, for instance by combining neural networks with continuous logic and multi-criteria decision-making tools (Csiszár et al., 2020) leading to the definition of Logical Neuronal Networks (LONNs). Recently, this methodology has been applied to recommender systems in medicine, providing in this way the option to define the logical combination of a hierarchy of parameters (Ochoa et al., 2021). However, one limitation of this methodology is its inherent inflexibility, which could be responsible for low performance of this modeling methodology (Wang, 2021). In the medical field, the customers want to have control of the logical combinations, and the smaller and more well-defined as well as generalizable the network is, the easier it is to make it explainable to the customers, perhaps sacrificing in this way the precision of the network.

To explainability also belongs the possibility to handle inherent uncertainties from the data. The recording of health data is not prone to errors and uncertainties, starting from the inherent biological variability responsible for different physiological responses, to errors in the recording of diagnoses (which is about 63% in EHRs^1^), leading to a persistent uncertainty in the recorded data in EHRs. This implies that models should be essentially non-deterministic, and that the model must be stochastic, rather than flexible or “smarter” in its architecture. Thus, an explainable model must be able to tell the customer how good its precision is by informing the corresponding amount of uncertainty^2^. Indeed, explainable models not only concern the transparency of the model’s structure and the information flow, leading to a final prediction based on relatively simple and generalizable models (Occam’s razor), but also imply the possibility to generate different parallel models reflecting in this way this uncertainty.

To this end, we have combined LONNs with Bayesian Neural Networks (BNNs) (Lampinen and Vehtari, 2001) in a novel modeling methodology we have defined as BaLONNs. We have tested this novel approach for the prediction of the kind of heart failure (HF) and the expected therapy time (TL) of patients with diabetes, using a Pakistan Data Base from the UCI repository (Chicco and Jurman, 2020). Based on these results we thus aim to develop not only an improved explainable model, but also a human centered application that informs the customer when the model is “unsure” about a given prediction.

In the next section we introduce the methodology and modeling strategy. Thereafter we test the methodology and provide qualitative as well as validation results. Finally, we discuss the implications of the introduced methodology and provide an outlook of the next research steps.

## Methodology

### Data extraction, feature engineering and data balancing methods

The data used in this analysis is composed of diabetic patients, some with heart insufficiency (Pakistan Database, from the UCI repository^3^). The corresponding attributes are listed below, where *V1* to *V10* are the inputs, and O1, as well as O2, are the model targets. Observe that the parameter “Time” is a metric of the total time (in months) that the patient has been treated.

Since we required a large population, we synthesized additional patients from the original database. Personal information, like the identification number (ID), age, and sex, were randomly generated. For the simulation of the distribution of diagnoses, we used the “synthpop” package^4^, basically using linear regression models for each parameter (Nowok et al., 2016).

From this data we can extract two main features:

- the kind of heart failure *HF* as a binary value representing patients with systolic heart failure (SHF, *HF = 1)* and heart failure with preserved ejection fraction (HFNEF, *HF = 0)*
- the therapy length *TL*, representing the time the patients are treated. In this last case, we transform the registered time τ, which is a natural number τ∈ℕ(number of months), into a discrete scale representing low (*TL = 0*, for 0 to 1 month), medium (*TL = {1,2}*, for 2 to 4 months), and a large (*TL = 3* for 5 and more months), expected therapy time.

While the first parameter *HF* is related to the kind of therapy a patient becomes, the second one, *TL*, is related to the quality of this therapy. Therefore, this one is a problem with two different tasks (TL, HF) and can be defined as a Multitask Learning (MLP) problem, where multiple tasks are simultaneously learned by a shared model.

In order to solve this problem, we opted to implement the model as a regression algorithm. *This seems a natural strategy because regression algorithms, by definition, have a notion of relative distance of target values, while classification algorithms usually do not* (Crawshaw, 2020).

Since these classes are essentially disbalanced, we require balancing methods to reduce bias in our modeling. To this end we implemented Synthetic Minority Oversampling TEchnique (SMOTE) to both *HF* and *TL* in order to create an oversampling of less frequent values. Despite its limitations, we selected this method because, according to Blagus et al., it is beneficial with low-dimensional data (Blagus and Lusa, 2013). In Figure 1 we present the original data distribution of *TL* before and after data balancing with SMOTE.

**Fig. 1.**
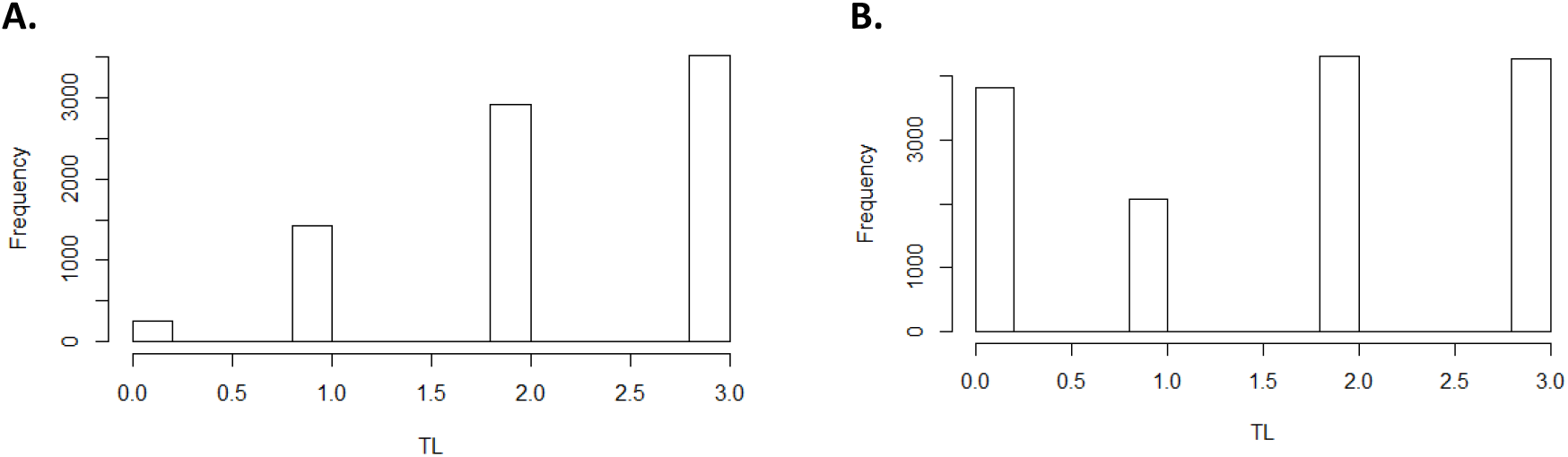
Distribution therapy lenght in the population in the original population (left) and the SMOTE balanced population (right)

From this data, 80% is used exclusively for model training and validation. The final 20% of the data is test data used to evaluate the quality of the model. For both, model training and quality evaluation, we normalized the input data.

### Baseline models

Our baseline model is essentially a deep-learning model with dense fully interconnected neuron layers implemented on Tensorflow.

The basic architecture of the baseline model is resumed in Table 2. Observe that in this model we have implemented rectified linear layers. Recent investigations have demonstrated that rectified linear functions are the most effective in representing data processing in neural networks^5^, particularly for networks with many layers (Urenda et al., 2020). Additionally, the ordinal classification problem of the therapy length is, based on Kramer et al., handled as a regression problem with an additional post-processing step (Kramer et al., 2001).

**Table 1.** Principal input and output parameters extracted from the HER of diabetic patients with heart insufficiency.

| Input / Output | Variable |  | Kind of parameter |
| --- | --- | --- | --- |
| - | ID |  | Character |
| V1 | Sex |  | Binary |
| V2 | Age |  | Real value |
| V3 | Creatinine_phosphokinase |  | Real value |
| V4 | Ejection_fraction |  | Real value |
| V5 | High blood pressure |  | Binary |
| V6 | Platelets |  | Real value |
| V7 | Serum_creatinine |  | Real value |
| V8 | Serum_sodium |  | Real value |
| V9 | Smoking |  | Binary |
| V10 | Anemia |  | Binary |
| O1 | Diagnose (kind of hearth insufficiency) | HF | Binary |
| O2 | Time (feedback period) | TL | Multiclass |

Based on this model we then modify part of the internal layers into fuzzy-Boolean units. The architecture has been selected in order to compare a “conventional” model against a modified model with fuzzy layers.

**Table 2.** Model parameters of baseline model

| Layer | # Units | Activation function |
| --- | --- | --- |
| Input Layer | 10 | Relu |
| #1 | 10 | Relu |
| #2 | 4 | Relu |
| #3 | 3 | Relu |
| #4 | 2 | sigmoid |

**Table 3.** Main model parameters used in the current implementation

|  | DNN | BNN | BaLONN |
| --- | --- | --- | --- |
| Optimizer | Adam | Adam | Adam |
| Learning Rate | 0.08 | 0.008 | 0.008 |
| Loss function | mse | negloglik | negloglik |
| Trainable Prior | - | True | True |
| Epochs | 400 | 300 | 300 |

### Bayesian Neural Networks (BNN)

In EHRs, errors arise in the way how data is registered, in part because the information in EHRs is in part manually curated, implying an epistemic uncertainty in the stored data^6^.

To correctly model this uncertainty, we implement models that *marginalizes over the distribution of parameters in order to make prediction*^7^ by implementing the weights in the neuronal network as a distribution defined as a Gaussian process. In this way, a trained model is not the result of the optimization of single parameters, but the optimization of the statistical distribution of these parameters. The same network with finitely many weights is known as a Bayesian neural network^8^. To this end, we aim to minimize the evidence lower bound of the network weights, which is defined as^9^^10^

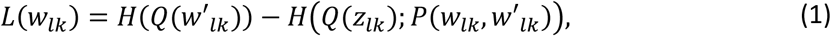

where *H*(*Q*(*w*^*′*^_*lk*_)) is the cross entropy defined as *H*(*Q*(*w*^*′*^_*lk*_)) = − ∑_z_ *Q*(*w*^*′*^_*lk*_) · *log* (*Q*(*w*^*′*^_*lk*_)), *Q* is a distribution over unobserved variables *w*^*′*^_*lk*_, in this case the prior, and *P*(*w*_*lk*_, *w*^*′*^_*lk*_) is the posterior of the distribution of observed data *w*_*lk*_, defined as a likelihood function. In this specific implementation the observed data *w*_*lk*_ are the neural-network’s weights of the layer *l* to the layer *k*, and *w*_*lk*_ is the estimated distribution for these weights. This definition is equivalent to minimizing the Kullback-Leiber divergence *D*_*KL*_(*Q* ∥ *P*) of the distributions *Q* and *P*.

*Thus, the network will be trained such that D*_*KL*_(*Q* ∥ *P*) → 0, *as well as maximize the probability of the data under the posterior weights*^11^: the model fit the actual achieve high *log likelihood*, while it stays close to the prior^12^. We define both *Q*(*w*^*′*^_*lk*_)[*σ*_*pr*_] and *P*(*w*_*lk*_, *w*^*′*^_*lk*_)[*σ*_*po*_] as Gaussian distributions, where *σ*_*pr*_ is the corresponding standard deviation of the prior, and *σ*_*po*_ is the standard deviation of the posterior. Finally, in all the probabilistic models the last layer delivers the result as a distribution, with its own standard deviation; in all the experiments we have fixed this standard deviation to 1.0. The optimization process for the model training was computed using the Adam method (Kingma and Ba, 2017).

This implementation has been performed on TensorFlow in R as well. The final trained model is an object containing the training functions for the distributions in the statistical layers. The final predictions are then obtained when a sample of models is computed, such that an individual model is the result of a specific set of weights for the links computed inside the probabilistic distribution. For this investigation, we have computed 1000 different models, which is enough to get a reasonable model sample.

These Bayes layers are then used to evaluate the effect of epistemic uncertainty in two models

- BNN: Bayesian Neural Networks (stochastic baseline model)
- BaLONN: Bayesian LONN

### LONNs and Bayesian Layers

The application of LONNs for the evaluation of medical data has been described by Ochoa et. al. (Ochoa et al., 2021), *whereby single layers are replaced by frozen weights and bias representing logical operations. Here, a single Perceptron in the NN network is activated by so-called Squashing activation functions, a differentiable and, parametric family of functions that satisfy natural invariance requirements and contain rectified linear units as a particular case (Urenda et al., 2020a; Zeltner et al., 2020). These activation functions are in this framework defined as follows*

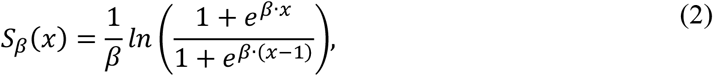

where *β* is a real nonzero value that must be adjusted to let the model be convergent. Thus, the Perceptron in the neural networks’ hidden layers can model a threshold-based nilpotent operator (Csiszár et al., 2020a, 2020b): a conjunction, a disjunction, or even an aggregative operator. *This means that the weights of the first layer are to be learned, while the hidden layers of the pre-designed neural block, work as logical operators with frozen weights and biases*.

**Figure 1.**
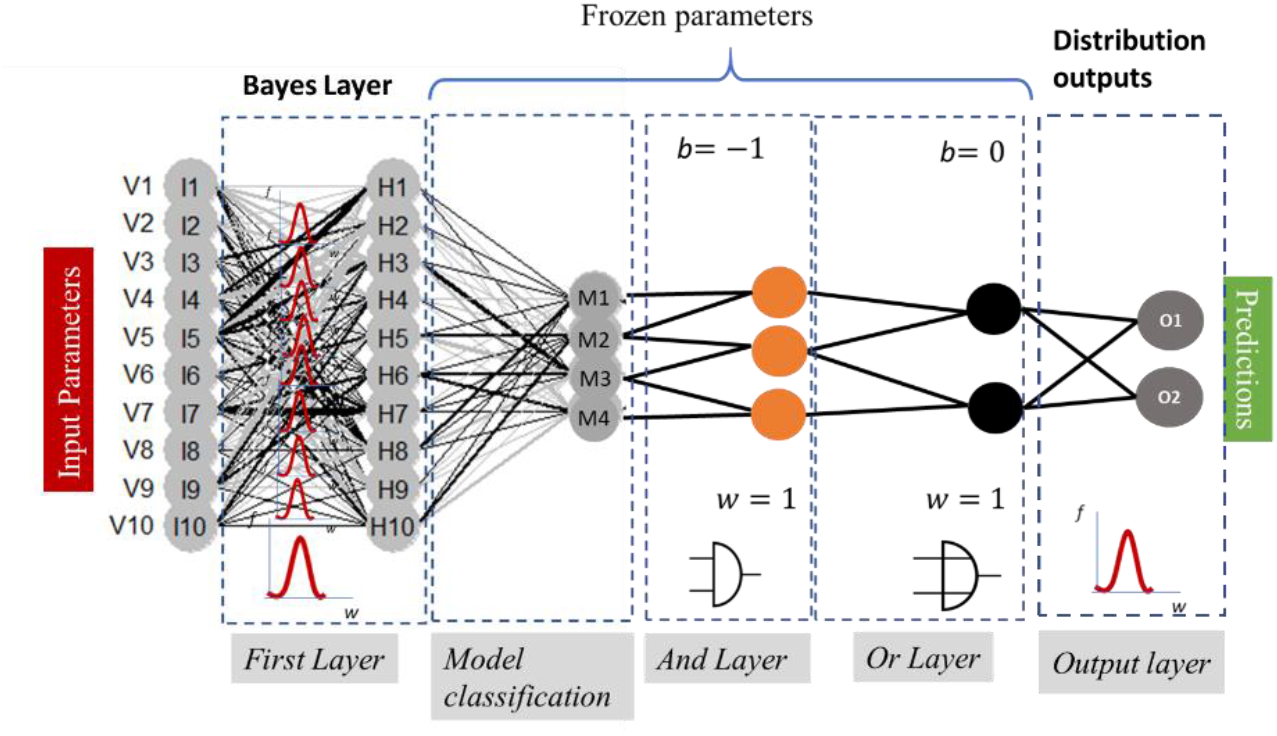
ProbLONNs, with the introduction of distributed weights in the initial layer representing the weight distribution according to the Bayes theorem. The number of neurons in the deep layers is defined according to the parameters presented in table 2

Thus, the baseline model is modified by freezing the weights and biases in the layers where we represent the logical switches.

**Table 2.** Some examples of logical operators and their corresponding implementation (Csiszár et al., 2020c)

| Logical operation | $w_{ij}$ | $b_i$ |
| --- | --- | --- |
| AND | 1 | -1 |
| OR | 1 | 0 |
| NOT (x) | -1 | 1 |
| NOT (y) | -1 | 1 |
| Not (x) and Not (x) | -1 | 1 |

This definition implies that the number of trainable parameters for the weight estimation gets reduced, implying also a reduction of the degrees of freedom of the model. Our implemented **Logic-Operator neural network (LONN)** is thus a method to simulate the cognitive logical thinking process (Figure 2), considering that this logic is joined by fuzziness, i.e., logical operations are not exact but essentially fuzzy due to the implemented continuous-valued operators.

**Figure 2.**
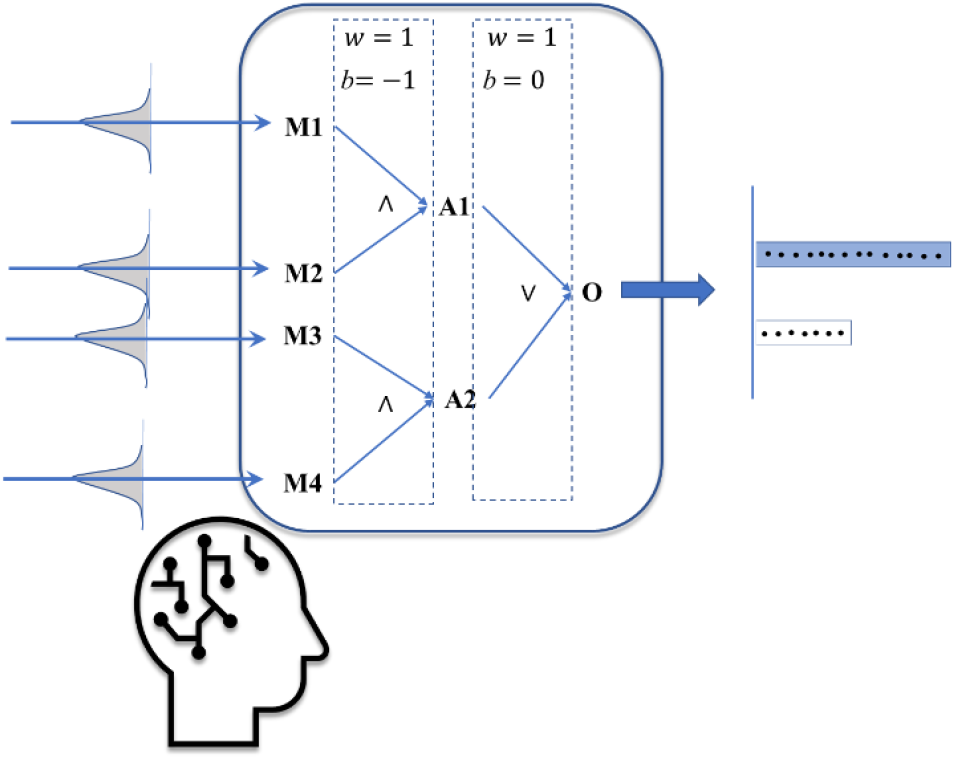
The outputs of the model are in some cases non deterministic, but ambiguous

By introducing probabilistic layers, we are not only simulating the possible structure of thinking processes, but also its inherent uncertainty. Therefore, the outputs from the model are not intended to present exact solutions and outcomes; rather, these outputs present both a plausible prediction of an outcome and its corresponding uncertainty.

### Results and Model Validation with Ambiguity

Based on the previous definitions we performed different computations using the meta-parameters for each model listed below.

The fact that we are dealing with uncertainties implies that the final result is non-deterministic, i.e., the outputs can consist of a sample of several plausible outputs. For instance, for some patients the evaluation of the expected therapy time can be a range (for instance, the expectative is that the patient will get a medium to long therapy time TL), and not an exact outcome (either a medium OR a long TL). This ambivalent scenario is much more realistic in a real clinical field than a pure deterministic case. For this reason, the validation must be performed accounting distributed outputs.

To compute the final validation and its corresponding statistics, we define the true prediction for each patient *i*(*Pr*_*i*_) for both HF and TL as the statistical value that best matches the expected target value. For the computation of *Pr*_*i*_ we perform a micro statistic, where the average of the predicted values *f*_*ij*_ is compared to the target value *O*_*i*_. If the distance between the average and the target lies below a tolerance value, then this state will be considered as a true prediction *Pr*_*i*_ = 1

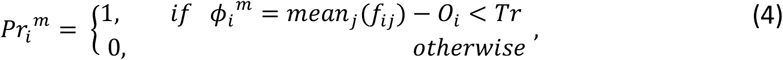

where *mean*_*j*_ is the mean value estimated over the index *j*. In our validation we opt to use micro averages^13^, i.e., biased by class frequency.

Since TL is a multi-label parameter (no false positive or negative as a binary state), and we want to validate HF and TL, we will only make an estimation of the quality of the validation using the metrics in equations 3 and 4. Furthermore, we defined the threshold values with a light bias in order to accept the output of two plausible values, according to the idea represented in figure 2:

- For *Tr*_*HF*_ we consider the prediction of a binary value between 0 and 1, such that 0 < *ϕ* < 1.0, such that *Tr*_*HF*_ = 0.7is a reasonable definition to sample a majority of one of the two binary values.
- For *Tr*_*TL*_ we are evaluating the distance between different states between 0 and 3. Contrary to the binary prediction (which can be either 0 or 1) we can accept predictions with more than two values, for instance 2 and 3 (expectative of a medium to large therapy time), such that 0 < *ϕ* < 2, such that *Tr*_*TL*_ = 1.2 is a reasonable definition to sample at least two plausible model outputs.

In a nutshell, *Tr* is a parameter to decide how many simultaneous states from the output *f*_*ij*_ can be accepted and is the accepted degree of ambiguity of the model (this fact is also visualized in figure 4).

Based on these two definitions, we can then compute the precision as a function of *TP*_*i*_ as

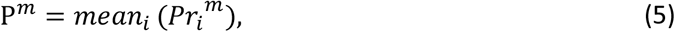

while the total error was estimated as

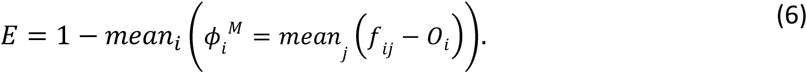

In order to test the correct functionality of the models, we perform a first inspection of the distribution of the predictions. For the baseline model (NNs) we directly represent the predicted distributions, while for the BNN we compute the mean value of the distributions. With this first general result we discover, first, that the model has variability and, second, can reproduce the dynamics of the test data. Furthermore, it is necessary to analyze if the variability, in particular in the BNNs, is generated by the model structure, as well as the information integration, and not by the stochastics of the probabilistic layers.

These first results demonstrate that the BNNs tend to make a different sampling in the predicted values: since this distribution samples the absolute values (low tolerance, do not accept two simultaneous values, but round the average output) the final distribution strongly differs from the target (Figure 3). Therefore, the modeling of the uncertainty as well as the acceptance of the ambiguity of the outputs, can influence the overall distribution of the prediction.

**Figure 3.**
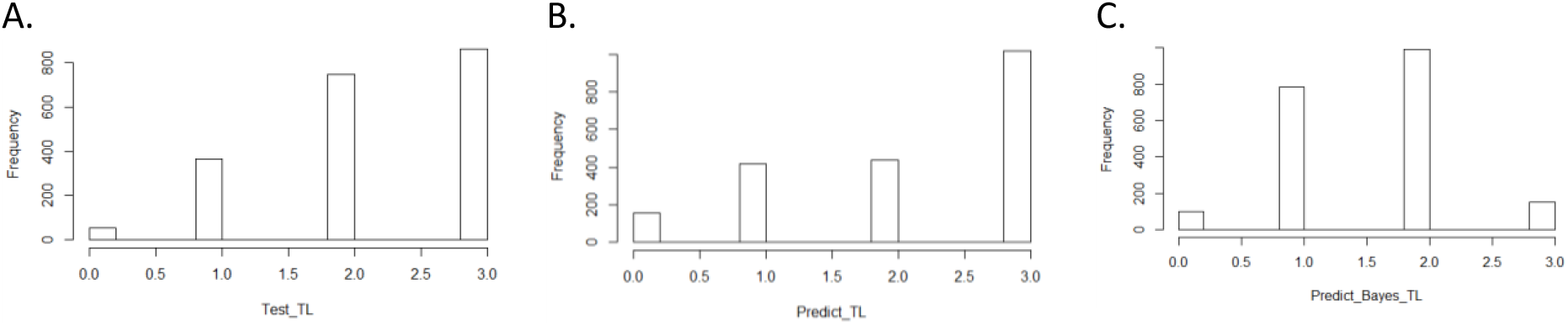
Distribution of the therapy length (TL) for the reference data (A), the prediction computed with the baseline model (B), and the BNNs (C)

We first explore the performance of the BNNs in order to better understand the behavior of the models, before we implement this concept to BaLONNs. To better assess the quality of the predictions we require a detailed validation, also considering that outputs can be ambiguous, as has been suggested in Figure 2. Thus, for the prognosis of TL of a single patient there is a chance to get for instance a short to middle expected TL; but the distribution shows a larger weight to one of the prognoses, as is shown in Figure 4. This implies that the main expectation is a short TL, but with a low chance to have a middle TL. The representation of this expectation for a patient population implies that the predicted outcomes are not simply a single prediction, but a distribution that can be represented for instance with a boxplot. In Figure 4, we deploy this boxplot for a small fraction of the patient population and measure the distance to target values to validate the model. The color keys represent the quality of the validation, with green as a perfect validation, blue as a prediction where the target value lies inside the distribution of the predictions, and red as an incorrect prediction, where the target lies outside the distribution of the computed predictions (Figure 4).

**Figure 4.**
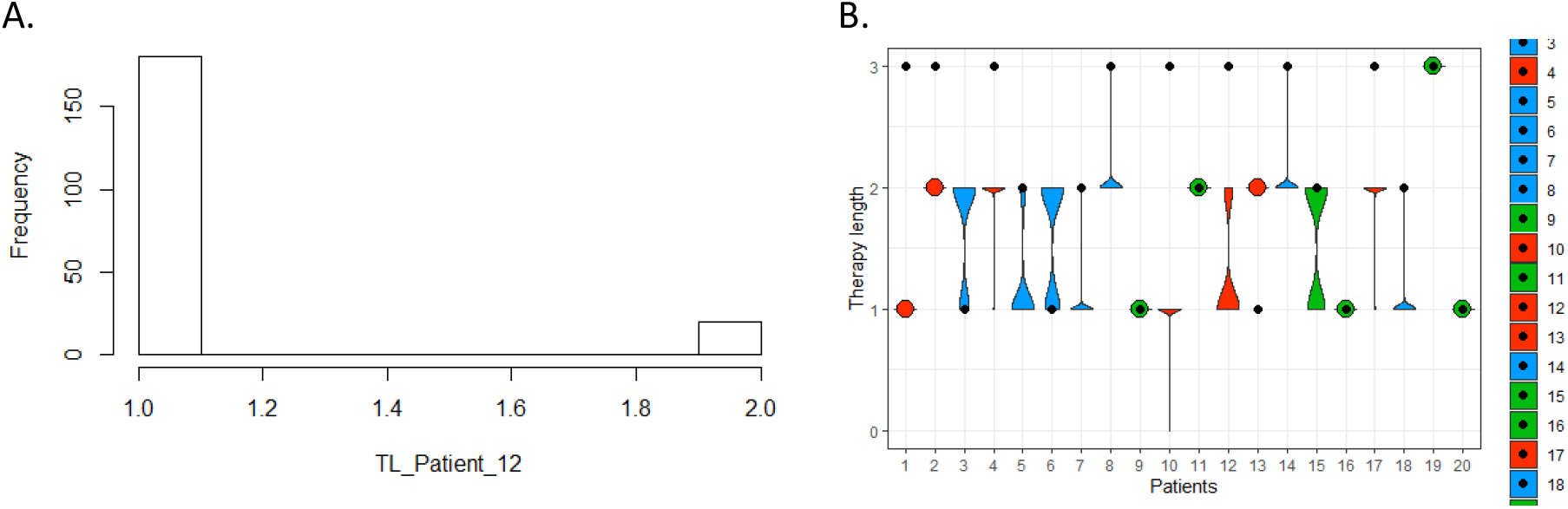
Precision of the prognosis of the therapy length for one patient with an ambiguous output (left), and for a cohort (20 patients) in the whole population with validation results: green is a perfect prognosis, blue is a by-prognosis that is still fulfilled inside the uncertainty of the prediction, and red is an off-prognosis. The black points are the targets.

The computation of the error considers the fact that both observables, therapy length and class of heart failure, are distributed as well. Finally, the results can be either represented using box plots (as in Figure 4) or with violin plots (Figure 5).

**Figure 5.**
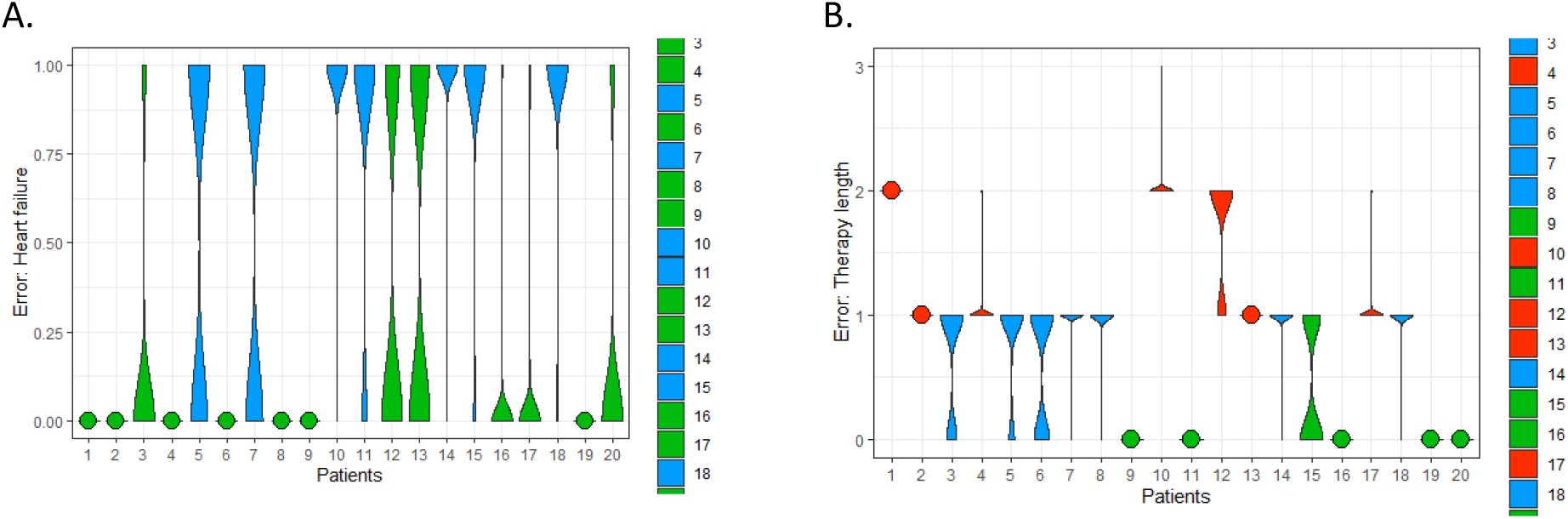
Predicted error of the predictions of HF (A, left) and TL (B, right)

These kinds of visualizations are useful ways to qualitatively understand how the implemented models are working and visualize when a model is relatively confident (green) or unsure (blue) about its prognosis, also depending on the standard deviation of the distribution. Of course, this has an implication for the final validation: with this implementation, we were able to reduce the error to *24.48* % and *21.24 %*, representing an improvement of about$*3%*$ in respect to pure deterministic models (total error of baseline model was 37.7%). But this error reduction has been obtained because we account predictions “in blue”, i.e., we are accepting Off-predictions, where the targets lie inside the predicted distribution. Otherwise, the computed predictions of the BNNs only considering the true positives/negatives could deliver a much higher prediction error (see Figure 5).

To get a better insight into the performance of the model, we assessed the model sensitivity to the standard deviation of the prior, *σ*_*pr*_, and of the posterior, *σ*_*po*_ and in this way analyzed the role of the distribution of the prior/posterior in the final result (Figure 6). This iteration is also a test about the robustness of the model by changing conditions or inputs, which is equivalent to performing adversarial tests on trained networks. The computation of these results was computationally intensive (about 2 hours) on a normal desktop. This implies that any parameter optimization considering this variation could require large computational resources in case we deal with larger datasets.

**Figure 6.**
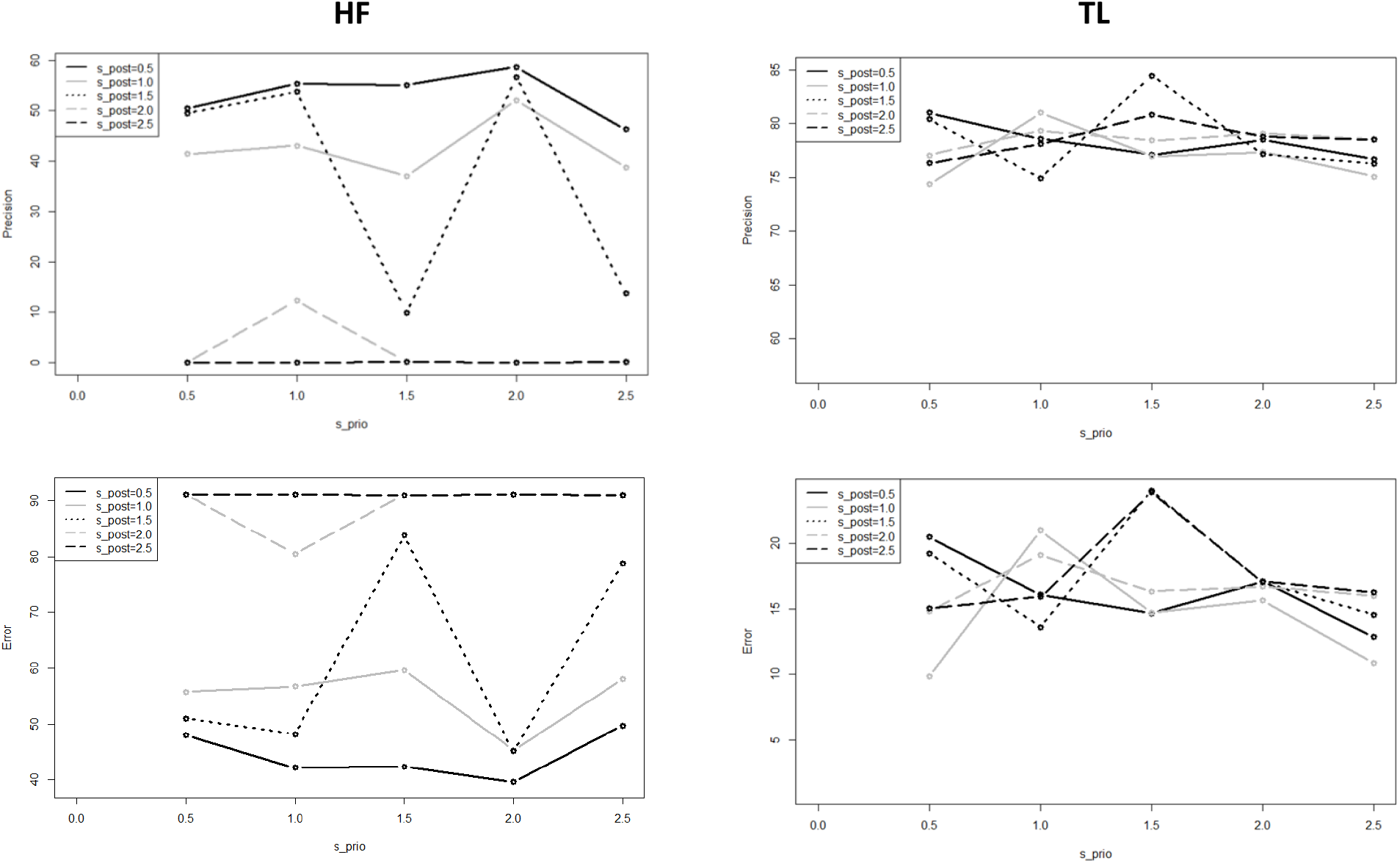
Precision of the predicted HF and TL using BNNs for different standard deviations; σ_pr_ (x axis), and σ_po_ (different shapes)

In this test, we observe for almost all the values a high model sensitivity on the prior and posterior standard variations. Also, the precision obtained for *HF* is smaller than the precision obtained for TL, due that *HF* is a binary output and hat itself fewer degrees of freedom than the multilabel output TL; we can understand the behavior of this state like a physical binary state that gets polarized beyond a critical value in the fluctuations of the system, represented by *σ*_*po*_. Thus, we observe that around *σ*_*po*_ = 1.5 (and *σ*_*pr*_*∈*[0.5, 2.5]) the *HF* states get unstable, and for *σ*_*po*_ > 1.5 the overall precision by *HF* dramatically decreases^14^ (see Figure 6).

Since the standard deviation influences the distribution of the weights in the model, it is to assume that this definition must reflect the distribution of the uncertainty in the training dataset. Otherwise, the model follows its own dynamics, deteriorating its ability to fit experimental targets.

From these results, we conclude that one plausible combination is *σ*_*pr*_ = 1.5 and *σ*_*po*_ = 1.0.

When we have tested again the model sensitivity for the BaLONNs we have observed that there is a lower fluctuation of the precision/error values for *σ*_*pr*_*∈*[0.5, 2.5]) and *σ*_*po*_ as control parameter (Figure 7). Like a physical system, by freezing the layers we are reducing the degrees of freedom of the network, reducing in this way the stochasticity, and in general the instability of the trained network. Also, in this case, we have observed that the *HF* precision is much lower than *TL*. Different from the BNNs, BaLONNs do not accept prior layers as trainable, which probably contributes to the deterioration of the HF validation.

**Figure 7.**
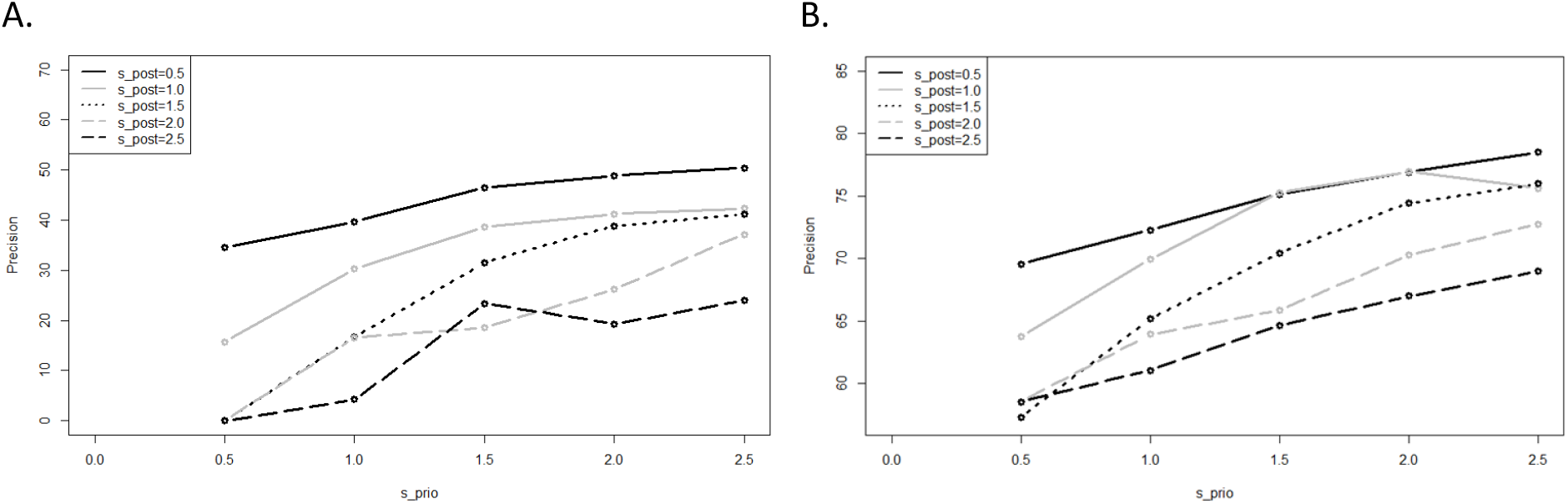
Computed precision of the BaLONNs for HF (left) and TL (right) and different standard deviations; σ_pr_(x axis), and σ_po_(different shapes)

We obtain acceptable validations for *σ*_*pr*_ = 1.5 and *σ*_*po*_ = 1.0. We finally have computed the final predictions in a single computation. For this computation we have considered a *HF* tolerance of 0.6 (for the loop the *HF* tolerance was 0.6) and made an average of over 1000 models (see table 4).

**Table 4.** Validation results for the baseline model, BNNs and BaLONNs. For the stochastic models we have averaged over 1000 different models.

|  | NN-Baseline Model |  |  | BNN |  |  | BaLONN |  |
| --- | --- | --- | --- | --- | --- | --- | --- | --- |
|  | HF | TL |  | HF | TL |  | HF | TL |
| ***E* (%)** | 60.9 | 4.73 |  | 43.5 | 15.13 |  | 43.59 | 12 |
| **Tot *E* (%)** | 32.81 |  |  | 29.31 |  |  | 27.79 |  |
| ***Pr* (%)** | 42.05 | 55.08 |  | 65.69 | 80.15 |  | 61.42 | 77.14 |
| **Tot.*Pr* (%)** | 48.56 |  |  | 72.92 |  |  | 69.28 |  |

The modeling of the epistemic uncertainty increases the precision of the modeling, simply because we are accepting ambiguous results as true predictions inside the tolerance range, while the base model is trying to make an exact and predictive prediction based on the training data. This effect reflects the fact that the inherent stochastics play a relevant role at the moment we want to make a prediction.

Furthermore, we have observed that the validation of *HF* using BaLONNs does not get the same quality as a fully trainable model, despite the mean error lying below the mean error of BNNs. Due to the fact that the logic gates are frozen /non-trainable parameters, the whole model has fewer degrees of freedom, limiting the possibility that the model converges into the target values. In this case we face again the typical tradeoff problem: either to accept dealing with interpretable models with less accuracy, or to have an accurate model with less interpretability.

We have also observed that while the precision of HF gets increased by adjusting the model’s hyperparameters, the TL’s precision decreases. The integration of these parameters in a single model is to some degree problematic, firstly considering that both parameters have a different number of states, and secondly, because their coupling is weak, i.e., TL depends on HF as well as additional parameters that are not integrated into the model. This was in particular evident in the NN-baseline model. Despite this, we could find a good tradeoff in the parameter selection to deliver an acceptable accuracy for both parameters.

## Discussion

The definition of explainability usually focuses on methods to make the outcomes from black-box models, like DNNs, interpretable. For instance, LIME and SHAP explore and use the property of local explainability to build surrogate models to black-box machine learning models to provide them with interpretability^15^. In our approach, we have opted for a different strategy by implementing nodes in the DNN model that can be defined as logical operators, LONN, which are interpretable. By doing this, we set up a hierarchy of parameters in the input layers that can then be logically combined, without requiring additional surrogate models.

However, the problem of explainability does not only focus on the interpretation of how information is integrated and how the corresponding outputs are computed depending on the interlinking of the inputs. It also concerns the implicit handling of uncertainty in the data. And this factor is particularly relevant in the medical field, where biological variability has an influence on the development of a disease. Thus, while for one patient a specific diagnosis or outcome after therapy can be clearly established, for another patient there can be an ambiguity that cannot be simply ignored.

Therefore, interpretability must consider not only the information flow and output computation in a model, but also the fact that in certain cases it is not possible to estimate an exact value. Thus, the kind of models considering epistemic uncertainty and explainability, like our BaLONNs, are somehow humble assistants that inform the customer when they cannot behave in a deterministic way, informing when they are unsure about the prediction they are delivering.

We consider that the inclusion of this kind of ambiguity in assistant systems, like recommender or expert systems in medicine, is an important step, also in the design of human-centered solutions. Several studies have shown that trusting *digital systems beyond their capacity and functionality can present high actual costs, as well as more nuanced effects of compromising organizational integrity and personal security* (Hardré, 2016). This is, in particular, relevant in the medical field, where the pressure in daily business might lead to strongly rely on automatic systems (Goddard et al., 2012), which, despite being well-validated, could lead to wrong decisions when customers assume that the outputs are precise when in reality are wrong.

Our novel approach is essentially an integration of LONNs with probabilistic layers. Our preliminary results have demonstrated that this approach can fulfill its goal: we are able to get acceptable validation values as well as an acceptable precision in the prediction of the two parameters used in this study, namely the prediction of the kind of heart insufficiency (HF) and the expected therapy time of the patients (TL). However, the precision of the model lies below a conventional deep learning model (which can be much more precise if we increase the number of layers). As we have found in a previous study, the explainability implies a tradeoff between model performance and precision: while explainable models are often small and constrained to few elements to keep them explainable, the expected precision is in this case much lower than for classical black-box models^16^. The customer must decide at the end how much precision she wants to sacrifice in exchange for a simple model.

The potential next step in this work is to introduce a level of stochasticity not only in the extreme layers but also in the logical switches, as well as in the number of these switches. In this way, we generate different explainable models, assuming that not only one but a family of explainable models is able to model the data. An additional analysis, not performed in this study, is the relation between stochasticity and minority oversampling using SMOTE, i.e., if by implementing SMOTE the underlying epistemic uncertainty of the training datasets gets distorted. Finally, the handling of the stochasticity could be computationally intensive when large datasets are analyzed; perhaps alternative methods, like quantum computing, could be helpful to generate more efficient stochastic BaLONN models from large datasets.

## Conclusion

In this work, we have presented a novel modeling technique that combines explainable model architectures based on logic neural networks (LONNs) and Bayesian methods in a single BaLONN model. In this way, we also are helping to improve user-centered solutions that inform the user when they are unsure about a prediction, in this case, the prediction of a therapy depending on physiological parameters. This contrasts with common solutions that nudge customers to accept the model prediction.

We have demonstrated that our model reaches an acceptable precision of 69%, considering that some of the true predictions are ambiguous but lie in the tolerance range to accept the prediction. This precision lies below the 70% precision of Bayesian neural networks. However, we think that the slight reduction of the precision is an acceptable tradeoff considering that the architecture of the NN is composed of few neurons, and that some of these neurons are non-trainable.

This result is thus a relevant basis for the development of assistant and expert methods, for instance, recommender systems, in critical fields like medicine where the customer’s work should not be fully automated, and his expertise is continuously required.

## Data Availability

All data produced are available online at
https://archive.ics.uci.edu/ml/datasets/Heart+failure+clinical+records

## Acknowledgments

We want to thank Martin Grundman and Thomas Schimper for their ideas and critical feedback in the development of these concepts. The project no. 2019-2.1.11-TÉT-2020-00217 has been implemented with the support provided from the National Research, Development and Innovation Fund of Hungary, financed under the 2019-2.1.11-TÉT-2020-00217 funding scheme.

## Footnotes

1 https://healthinfoservice.com/most-common-icd-10-error-codes/

2 https://www.heise.de/hintergrund/Wie-eingebautes-Misstrauen-KI-Systeme-sicherer-machen-kann-6049154.html?wt_mc=rss.red.ho.ho.atom.beitrag.beitrag

3 The data has been obtained from the [uci repository](https://archive.ics.uci.edu/ml/datasets/Heart+failure+clinical+records)

4 https://www.r-bloggers.com/generating-synthetic-data-sets-with-synthpop-in-r/

5 https://scholarworks.utep.edu/cgi/viewcontent.cgi?article=2170&context=cs_techrep

6 In case of evaluation of time series, for instance in ECGs, we expect to find aleatoric uncertainty

7 https://cedar.buffalo.edu/~srihari/CSE574/Chap5/Chap5.7-BayesianNeuralNetworks.pdf

8 https://blogs.rstudio.com/ai/posts/2019-06-05-uncertainty-estimates-tfprobability/

9 https://en.wikipedia.org/wiki/Evidence_lower_bound

10 https://xyang35.github.io/2017/04/14/variational-lower-bound/

11 https://blogs.rstudio.com/ai/posts/2019-11-07-tfp-cran/

12 https://blogs.rstudio.com/ai/posts/2019-06-05-uncertainty-estimates-tfprobability/

13 https://stats.stackexchange.com/questions/156923/should-i-make-decisions-based-on-micro-averaged-or-macro-averaged-evaluation-mea

14 This observation resembles a phase transition, where *σ*_*po*_ has influence on the HF states. Up to a *σ*_*po*_>2.5all the states HF are frozen, i.e., the model has no variability, and the outputs are fixed to a single state.

15 https://towardsdatascience.com/idea-behind-lime-and-shap-b603d35d34eb

16 https://www.interpretable.ai/interpretability/what/

